# Placental biomarkers in second trimester maternal serum are associated with postpartum hemorrhage: a secondary analysis of the NuMoM2b dataset

**DOI:** 10.1101/2024.12.30.24319779

**Authors:** Julienne N. Rutherford, Erin K. George, Elise N. Erickson

## Abstract

Postpartum hemorrhage (PPH) is the leading cause of maternal mortality worldwide, which is often attributed to retained placenta (RP) after delivery. There are no biomarkers currently used to predict a risk of developing RP/PPH prior to labor. The objective of this study was to determine relationships between placental biomarkers measured in the first and second trimesters and proxy measures of postpartum blood loss relative to preeclampsia status in the Nulliparous Pregnancy Outcomes Study: Monitoring Mothers-to-Be (nuMoM2b) dataset. 2,192 participants had placental analytes drawn during the first and second trimesters (9-13 and 16-22 weeks gestation, respectively); the outcome was a composite of retained placenta and/or PPH requiring blood transfusion (RP/PPH). Using Kruskal-Wallis tests, median differences in levels of soluble fms-like tyrosine kinase-1 (sFlt-1), placental growth factor (PlGF), sFlt-1/PlGF ratio, soluble endoglin (sEng), beta subunit of human chorionic gonadotropin (β-hCG), inhibin A (INHA), and pregnancy-associated protein-A (PAPP-A) were assessed between women with (n=67) and without (n=2125) RP/PPH overall and stratified by preeclampsia status. Women with RP/PPH had significantly higher median levels of sEng, β-hCG, INHA, PAPP-A in the second trimester and sFlt-1was higher in both first and second trimesters, which was observed again when stratifying by preeclampsia status. Our findings indicate that biomarkers associated with angiogenesis, particularly when measured in the second trimester, are important targets for further study of RP and/or PPH pathophysiology and potential risk screening development.

## Introduction

Postpartum hemorrhage (PPH), currently defined by the World Health Organization as blood loss >500 mL for vaginal births and >1000 mL for cesarean births within the first 24 hours [1, 2], is the leading cause of maternal mortality worldwide, accounting for nearly a third of maternal deaths [3, 4]. The rate of PPH in the United States is approximately ∼3%, affecting ∼200,000 women every year, and rates have been rising for the last 30 years [5], [6] largely due to increased use of labor induction and previous cesarean birth [7]. Only about 10-40% of eventual PPH cases are predicted by current clinical risk-assessment tools [8-10]. Estimates suggest as many as 54-93% of deaths due to postpartum hemorrhage could be prevented [11]. Many PPH prognostic models have been proposed but none are ready for clinical application [12]. A novel way of thinking about PPH is necessary to discover pathways to early detection, treatment, and primary prevention. There are no biomarkers currently used to predict a risk of developing PPH prior to labor [13], largely because PPH, unlike preeclampsia, has not been considered as a disorder with roots in early pregnancy. In contrast, much progress has been made in the use of placental biomarkers for early detection of preeclampsia risk, due to a focus on earlier stages of placental development, particularly angiogenesis.

Several angiogenic biomarkers produced by the placenta have been identified as predictors of preeclampsia risk. Preeclampsia is a hypertensive disorder of pregnancy largely viewed as the consequence of impaired angiogenic and maternal endothelial dysfunction. Placental growth factor (PlGF) and soluble fms-like tyrosine kinase-1 (sFlt-1) are two trophoblast-derived molecules which play an important role in placentation and outcomes related to invasiveness and remodeling. For example, PlGF is a placenta-specific member of the vascular endothelial growth factor (VEGF) family involved in both vasculogenesis and angiogenesis [14]; PlGF rises during pregnancy, peaking around 26-30 weeks gestation and decreasing toward term [15]. sFlt-1 has antiangiogenic properties and is important in the regulation of blood vessel formation in diverse tissues, including the uterus and placenta [16, 17]. sFlt-1 binds the pro-angiogenic factors VEGF and PlGF. sFlt-1 disrupts endothelial function during the remodeling of the uterine vessels [18]. The role of sFlt-1 and PlGF in the etiology of preeclampsia and their diagnostic value have been extensively studied and validated [19-22]. For example, an sFlt-1/PlGF ratio >85 at 20-33 weeks gestation was strongly associated with developing preeclampsia [23]. Other placental biomarkers associated either directly or indirectly with angiogenic processes have been found to predict preeclampsia. Soluble endoglin (sEng) is also an anti-angiogenic biomarker that behaves in a manner similar to sFlt-1 [24]. Increased sEng in the second trimester is associated with increased preeclampsia risk [25, 26]. Pregnancy-associated protein-A (PAPP-A) is a metalloproteinase that activates insulin-like growth factors (IGFs)[27]. Reduced PAPP-A in the first trimester, but increased in the second trimester is associated with preeclampsia [28, 29] [25, 26, 28]. Inhibin A (INHA) is a glycoprotein produced by the placenta. INHA is involved in regulation of implantation and trophoblast proliferation; elevated second trimester INHA is associated with preeclampsia risk [30]. The beta subunit of human chorionic gonadotropin (β-hCG) has many functions in pregnancy but plays a role in angiogenesis through stimulation of sprout formation [31]; elevated second trimester β-hCG has been found to be associated with preeclampsia [32, 33] though a recent meta-analysis indicates the evidence is not yet strong [34].

It has been suggested that like preeclampsia, PPH might also be a consequence of abnormal placental development and angiogenesis [13, 35, 36], particularly because PPH is often associated with retained placenta. However, to date, there has not been robust systematic study of placental biomarkers related specifically to postpartum bleeding outcomes or delayed placental delivery. The objective of this study was to determine relationships between placental biomarkers measured in the first and second trimesters and proxy measures of postpartum blood loss publicly available in the Nulliparous Pregnancy Outcomes Study: Monitoring Mothers-to-Be (NuMoM2b) dataset.

## Methods

The analytic sample is derived from the NuMoM2b dataset, which consisted of 10,038 nulliparous participants with singleton pregnancies who gave birth in hospitals across the United States between 2010-2015 [37, 38]. NuMoM2b was a prospective cohort study; complete clinical data during pregnancy, labor, and birth were collected, as were biospecimens, including maternal serum for biomarker assays, the results of which are publicly available via the National Institute of Child Health and Human Development (NICHD) Data and Specimen Hub (DASH) [38]. For NuMom2B participants for whom there were placental biomarker results, the analysis was first limited to participants with at least five 0.5 ml aliquots of maternal serum available during Visits 1 (first trimester, at 6-14 weeks gestation) and 2 (second trimester, 16-22 weeks gestation). The second restriction was to only include participants who delivered at or after 20 weeks gestation and had pregnancy outcome data available. Finally, participants with one or more of the following adverse pregnancy outcomes were selected: 1) delivery prior to 37 weeks gestation; 2) pregnancy complicated by preeclampsia or eclampsia; 3) newborn birth weight considered small for gestation age (<5th percentile); and 4) stillborn birth. A total of 2,419 participants were ultimately selected for placental biomarker analysis: 1,509 who met the restrictions and 910 without any of these conditions as controls. A total of 1,389 cases and 851 controls agreed to release their data for research purposes. We then excluded 48 participants with fetal congenital anomalies, yielding a final sample of 2,192 participants. Because neither blood loss quantity nor general PPH diagnosis were recorded in the original data, we constructed a composite PPH outcome of “retained placenta” and/or “hemorrhage requiring blood transfusion” (RP/PPH) as a proxy for the likelihood of greater blood loss. We examined the absolute difference in sFlt-1, PlGF, sFlt-1/PlGF ratio, endoglin, and hCG levels between participants who had RP/PPH and those who did not at both timepoints. We then examined differences in biomarker levels based on whether participants in either group were also diagnosed with preeclampsia. Differences in biomarkers between groups were determined using a Kruskal-Wallis tests of median differences, which is robust to assumptions of normality. To boost rigor of the analytic approach and due to the exploratory nature of the biomarker comparisons, we used a p-value of 0.004 to determine statistical significance to account for multiple comparisons using a Bonferroni correction.

## Results

The demographic variables and pregnancy/labor/birth outcomes of the sample are described in Tables 1 and 2, respectively. Of the 2,192 participants in this sample, 67 experienced RP/PPH (3.1%) and 2,125 did not (96.9%); women with and without RP/PPH did not differ in any demographic variables (Table 1). Preeclampsia was diagnosed in 572 of the total participants (26.1%); participants with RP/PPH were significantly more likely to have preeclampsia (Table 2). Cesarean birth and instrumental delivery were also more common in the RP/PPH group.

**Table 1:**
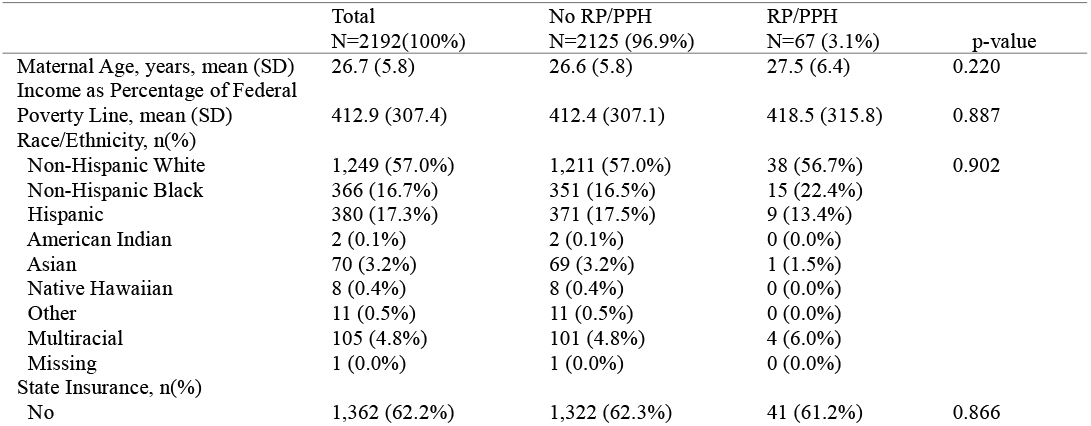
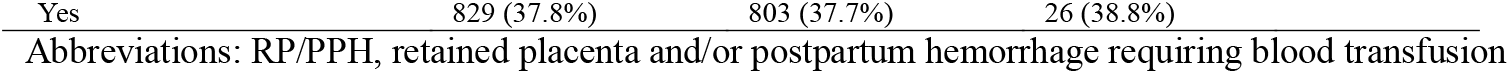
Sociodemographic Characteristics for NuMom2B Participants with Placental Analyte Results, by retained placenta/hemorrhage status (χ^2^& two-tailed t-test)

**Table 2:**
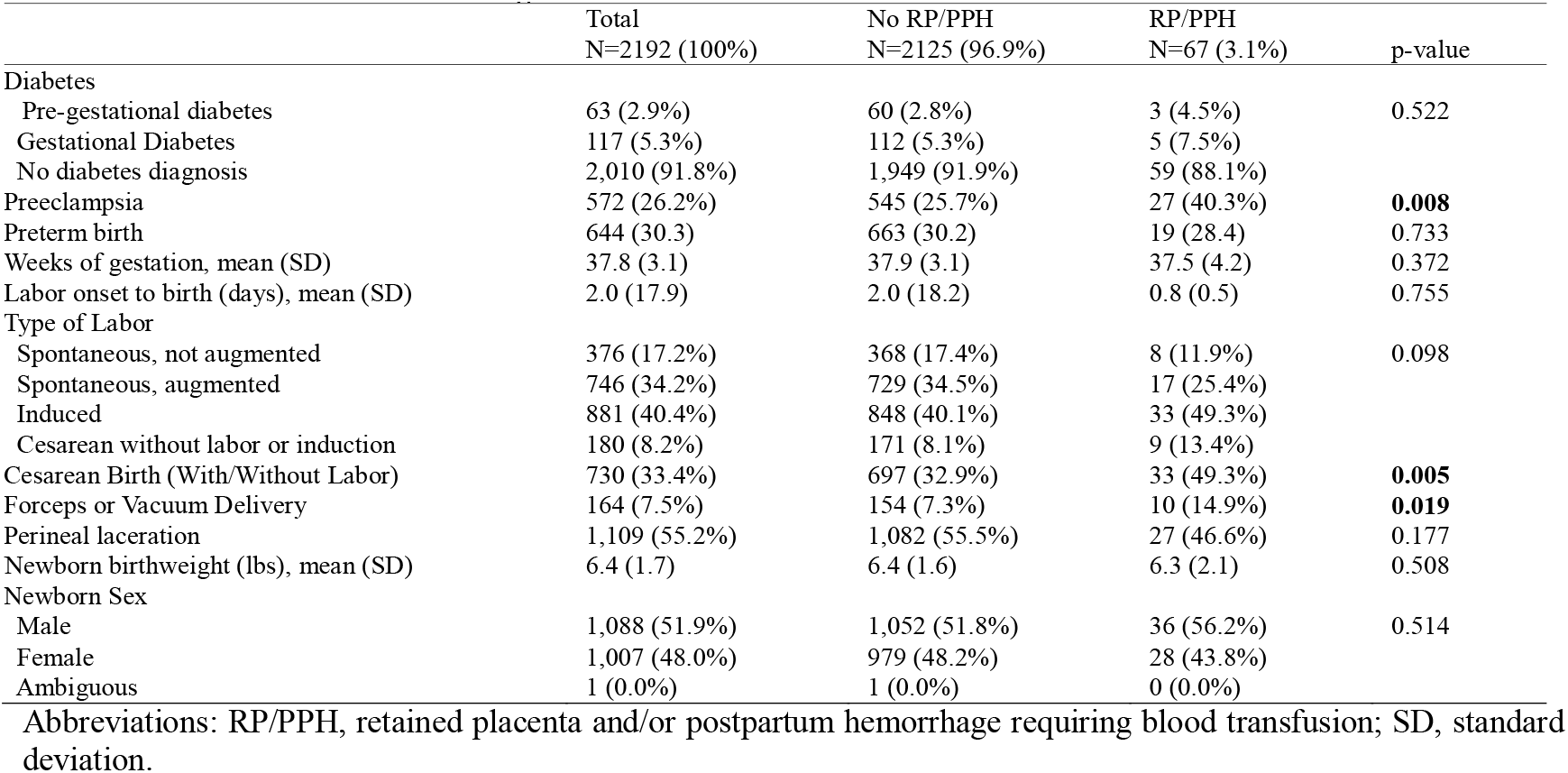
Pregnancy, Labor and Birth Outcomes for NuMom2B Participants with Placental Analyte Results, by retained placenta/hemorrhage status (χ^2^ & two-tailed t-test)

### Median second trimester angiogenic biomarkers were significantly elevated among women who experienced a retained placenta or postpartum hemorrhage at delivery

Across the sample, most of the median biomarker levels were significantly different between the two groups (RP/PPH & non) in the second trimester (Table 3). Participants who eventually developed RP/PPH had higher second trimester levels of sEng (p=0.004), β-hCG (p<0.001), INHA(p<0.001), PAPP-A (p<0.001) and sFlt-1 (p<0.001). The RP/PPH group also had a higher first trimester sFLT-1 (p<0.004). First trimester sFlt-1/PlGF ratio was also higher for the RP/PPH group. Other first trimester values were non-significant between the groups, and PIGF was not different at either time point.

**Table 3:** Placental Biomarker Results for NuMom2B Participants, by RP/PPH status (Kruskal-Wallis tests) Abbreviations: IQR, interquartile range; RP/PPH, retained placenta and/or postpartum hemorrhage requiring blood transfusion; sEng, soluble endoglin; β-hCG, human chorionic gonadotropin beta subunit; INHA, inhibin A; PAPP-A, pregnancy associate plasma protein A; PlGF, placental growth factor; sFlt-1, soluble fms-like tyrosine kinase-1; T1, trimester 1 (6-14 GW); T2, trimester 2 (16-22 GW)

| Biomarker<br>Median (IQR) | Timepoint<br>measured | Total<br>N=2192 (100%) | No RP/PPH<br>N=2125 (96.9%) | RP/PPH<br>N=67 (3.1%) | p value |
| --- | --- | --- | --- | --- | --- |
| sEng<br>(ng/mL) | T1 | 6.21 (1.95) | 6.20 (1.94) | 6.64 (2.16) | 0.041 |
|  | T2 | 5.40 (1.61) | **5.38 (1.60)** | **5.88 (1.88)** | **0.004** |
| β-hCG<br>(mIU/mL) | T1 | 19.89 (17.95) | 19.84 (17.92) | 22.46 (22.08) | 0.065 |
|  | T2 | 4.07 (3.89) | **4.05 (3.84)** | **5.97 (8.50)** | **<0.001** |
| INHA<br>(ng/mL) | T1 | 303.72 (210.04) | 303.04 (207.82) | 329.20 (258.78) | 0.082 |
|  | T2 | 204.16 (113.50) | **202.65 (111.77)** | **244.51 (137.58)** | **<0.001** |
| PAPP-A<br>(MU/L) | T1 | 990.68 (1507.90) | 977.75 (1479.87) | 1299.87 (1739.16) | 0.063 |
|  | T2 | 8810.17 (9439.11) | **8688.46 (9283.46)** | **12244.17 (12071.33)** | **<0.001** |
| PIGF<br>(pg/mL) | T1 | 41.17 (30.02) | 41.31 (29.97) | 37.45 (29.10) | 0.266 |
|  | T2 | 181.00 (150.77) | 181.41 (150.45) | 150.44 (181.11) | 0.590 |
| sFlt-1<br>(pg/mL) | T1 | 883.53 (511.29) | **875.95 (509.39)** | **1006.19 (621.84)** | **0.004** |
|  | T2 | 869.06 (573.54) | **864.10 (569.42)** | **1060.64 (858.13)** | **<0.001** |
| sFlt-1/PIGF<br>Ratio | T1 | 20.91 (18.11) | **20.78 (17.97)** | **27.59 (17.84)** | **0.002** |
|  | T2 | 4.66 (4.58) | 4.63 (4.55) | 6.87 (5.66) | 0.005 |

### Non-preeclamptic participants who had retained placenta or postpartum hemorrhage had significantly higher second trimester median levels of placental biomarkers than those who did not

We stratified the analysis by a diagnosis of preeclampsia and found the significant second trimester differences remained among nearly all the same biomarkers for the group without a co-morbid diagnosis of preeclampsia during the pregnancy (Table 4). Specifically, the median second trimester levels of β-hCG, INHA, PAPP-A and sFlt-1 were all higher among those who experienced a later RP/PPH.

**Table 4:** Placental Biomarker Results for NuMom2B Participants with and without preeclampsia, by RP/PPH status (Kruskal-Wallis tests)

|  |  | NO PREECLAMPSIA<br>N=1612 (73.9%) |  |  | PREECLAMPSIA<br>N=572 (26.1%) |  |  |
| --- | --- | --- | --- | --- | --- | --- | --- |
| Biomarker<br>Median<br>(IQR) | Time<br>Point | No RP/PPH<br>N=1572 (97.5%) | RP/PPH<br>N=40 (2.5%) | p value | No RP/PPH<br>N=545 (95.3%) | RP/PPH<br>N=27 (4.7%) | p value |
| sEng<br>(ng/mL) | T1 | 6.20 (1.90) | 6.83 (2.13) | 0.023 | 6.18 (2.14) | 6.38 (2.41) | 0.613 |
|  | T2 | 5.36 (1.53) | 5.67 (1.38) | 0.043 | 5.46 (1.84) | 6.16 (2.31) | 0.045 |
| β-hCG<br>(mIU/mL) | T1 | 20.12 (17.76) | 23.79 (19.01) | 0.084 | 18.74 (18.66) | 18.58 (28.91) | 0.277 |
|  | T2 | **4.10 (3.83)** | **5.90 (8.35)** | **0.002** | 3.86 (3.74) | 5.97 (12.24) | 0.009 |
| INHA<br>(ng/mL) | T1 | 302.94 (200.13) | 342.77 (271.86) | 0.095 | 301.19 (233.17) | 313.74 (262.55) | 0.488 |
|  | T2 | **199.89 (109.54)** | **254.95 (126.79)** | **0.003** | 213.62 (125.64) | 243.27 (160.95) | 0.164 |
| PAPP-A | T1 | 1007.14 (1500.53) | 1594.81 (1734.32) | 0.015 | 878.14 (1491.39) | 1008.16 (1344.54) | 0.890 |
| (MU/L) | T2 | **8868.42 (9179.38)** | **14812.99 (14402.51)** | **0.002** | 7995.78 (9810.36) | 10074.31 (8678.93) | 0.119 |
| PIGF | T1 | 42.34 (31.47) | 42.33 (27.61) | 0.927 | 39.18 (26.88) | 28.37 (30.25) | 0.081 |
| (pg/mL) | T2 | 190.06 (153.61) | 206.54 (302.51) | 0.503 | 162.64 (132.99) | 102.25 (166.75) | 0.142 |
| sFlt-1 | T1 | 887.95 (505.48) | 1044.02 (555.92) | 0.012 | 838.31 (521.06) | 950.41 (667.86) | 0.085 |
| (pg/mL) | T2 | **872.54 (580.30)** | **1174.96 (878.60)** | **<0.001** | 811.29 (559.93) | 918.89 (857.68) | 0.109 |
| sFlt-1/PIGF | T1 | 20.73 (17.67) | 24.59 (14.02) | 0.093 | 20.85 (18.26) | 32.04 (26.64) | 0.008 |
|  | T2 | 4.53 (4.10) | 6.73 (5.65) | 0.088 | 4.88 (5.69) | 7.18 (6.15) | 0.026 |

Abbreviations: RP/PPH, retained placenta and/or postpartum hemorrhage requiring blood transfusion; sEng, soluble endoglin; β-hCG, human chorionic gonadotropin beta subunit; INHA, inhibin A; PAPP-A, pregnancy associate plasma protein A; PIGF, placental growth factor; sFlt-1, soluble fms-like tyrosine kinase-1; T1, trimester 1 (6-14 GW); T2, trimester 2 (16-22 GW)

### Degree of change in biomarker levels differed in participants who experienced retained placenta or postpartum hemorrhage according to preeclampsia status

Figure 1 shows the Kruskal-Wallis median tests for differences in the percentage change in biomarkers between T1 and T2, for women who experienced RP/PPH stratified by preeclampsia. Preeclamptic women with RP/PPH had a significantly greater increase in PAPP-A and decrease in sEng from T1 to T2. Non-preeclamptic women with RP/PPH had a significantly greater increase in sFlt-1 from T1 to T2. The percentage change over time in INHA, β-hCG, and sFlt-1/PlGF ratio for women did not differ according to preeclampsia status.

**Figure 1:**
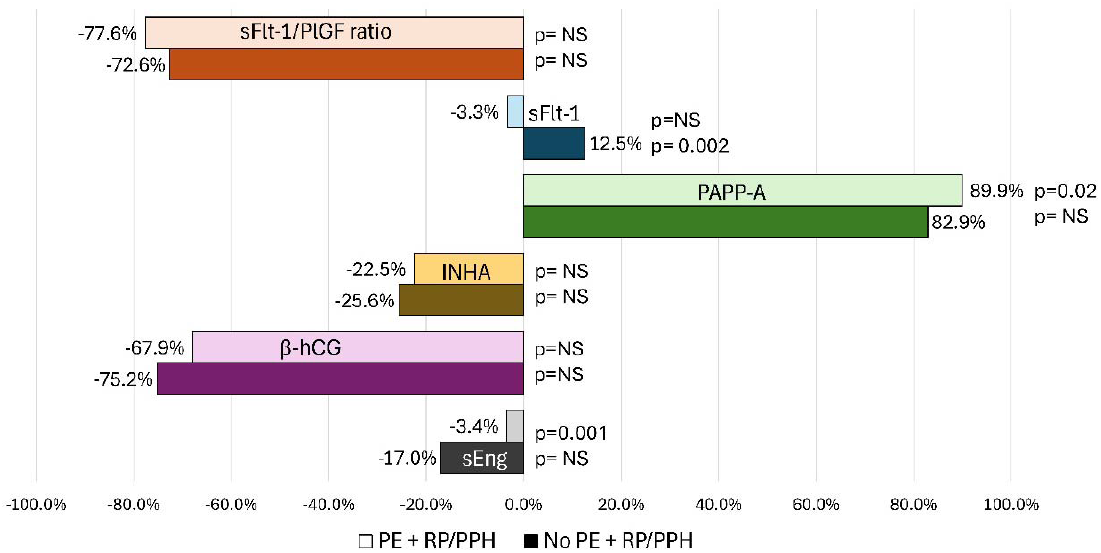
Percentage change from first to second trimester for sEng, β-hCG, INHA, sFlt-1, and the sFlt-1/PlGF ratio in women with RP/PPH, by preeclampsia status. Pale bars refer to preeclamptic women with RP/PPH; dark bars refer to non-preeclamptic women with RP/PPH. Abbreviations: RP/PPH, retained placenta and/or postpartum hemorrhage requiring blood transfusion; sEng, soluble endoglin; β-hCG, human chorionic gonadotropin beta subunit; INHA, inhibin A; PAPP-A, pregnancy associate plasma protein A; PlGF, placental growth factor; sFlt-1, soluble fms-like tyrosine kinase-1; T1, trimester 1 (6-14 GW); T2, trimester 2 (16-22 GW)

## Discussion

We have identified several biomarkers, all measured within the first 22 weeks gestation of gestation, that are significantly associated with postpartum hemorrhage variables. This is the first time that this many biomarkers have been studied together in a single sample to determine their relationship specifically to postpartum hemorrhage. Few studies have examined the relationship between individual placental biomarkers and postpartum bleeding on its own, but our findings are broadly consistent with the existing literature and provide strong rationale for further study. We found that sFlt-1 was significantly elevated in both trimesters for women who went on to develop RP/PPH. In 392 pregnancies not complicated by preeclampsia, Eskild et al. (2010) found that in normotensive women who went on to experience excessive blood loss (defined as >500 mL) compared to those who did not, sFlt-1was higher in all three trimesters, but significant only in the second trimester, and that the increase in sFlt-1 during pregnancy was greater for those who eventually experienced excessive blood loss [39]. Similarly, we found that while sFlt-1 did increase from the first to the second trimester for all participants, the size of the increase was larger for those who went on to experience RP/PPH. We found no associations between PlGF at either time point and RP/PPH, either in the sample overall or between preeclampsia groups. Ghosh et al. (2012) found that in preeclamptic women, PlGF<122 pg/mL at 22-24 weeks was significantly associated with PPH [40]. Our data come from samples collected at no later than 22 weeks; it is possible relationships with PlGF could emerge later in gestation.

We found that second trimester β-hCG was significantly elevated in the RP/PPH group. β-hCG levels measured 15-20 weeks gestation were strongly correlated with a later diagnosis of PPH (r=0.62, p<0.05) in a sample of 987 women [41]. In a recent retrospective cohort study of over 20,000 women, women with elevated β-hCG were more likely to experience PPH than those with normal β-hCG levels, though the effect size was small (0.05% vs. 0.03%, p=0.05) [42]. We found that second trimester inhibin-A was significantly higher for women who developed RP/PPH. While inhibin-A has been characterized in the context of preeclampsia we have found only one study that examined inhibin-A relative to postpartum hemorrhage. For example, one study found that women with elevated inhibin-A between 15 and 19 weeks gestation were more likely to develop preeclampsia; they did not look at PPH as an outcome [43]. Similarly, in a study of over 5000 pregnancies, high levels of inhibin-A measured at 15-20 weeks gestation were found to be correlated with preterm birth, preeclampsia, fetal growth restriction, and low birth weight; they did consider PPH as an outcome but did not find any relationship [44]. We found that second trimester PAPP-A was significantly elevated in women who go on to experience RP/PPH. There is scant literature on the relationship between PAPP-A and postpartum hemorrhage. In a recent study, first trimester PAPP-A multiple of the median (MoM) was significantly higher among women who experienced postpartum hemorrhage of less or greater than 1500 mL (MoM: 1.25±0.71, 1.9±0.69 respectively) than among women who did not experience hemorrhage (0.97±0.46, p<0.001) [45]. That study focused on samples collected during the first trimester antenatal aneuploidy screening thus they did not report second trimester PAPP-A levels.

Since angiogenic biomarkers associated with preeclampsia risk are relatively well-characterized, especially sFlt-1, and because PPH is more common for preeclamptic women generally and in the current sample, we stratified our analyses by preeclampsia status. We found that among women without preeclampsia, the relationships between the biomarkers and RP/PPH status were similar to those for the sample overall. Specifically, the biomarkers β-hCG, INHA, PAPP-A, and sFlt-1 were significantly elevated during the second trimester for these women. Conversely, for preeclamptic women, we found that none of the biomarkers significantly differed between the RP/PPH and no-RP/PPH groups. In other words, absolute levels of these biomarkers appear to effectively differentiate the potential for RP/PPH for women who do not have preeclampsia. We also looked at the magnitude of change in median biomarker levels from the first to second trimester in women who experienced RP/PPH, again stratified by preeclampsia. The magnitude of change in sFlt-1 was significantly greater for women without preeclampsia, while the change in sEng and PAPP-A was significantly greater for women with preeclampsia. These findings indicate that change in biomarkers, not just the absolute level of the analyte may a distinguishing feature for future PPH regardless of preeclampsia status or absolute biomarker levels. Together, our findings indicate that biomarkers associated with angiogenesis and other placental functions, particularly when measured in the second trimester, are important targets for further study of PPH pathophysiology and potential risk screening development. The second trimester is marked by tremendous placental angiogenic activity. The biomarker patterns we describe here may reflect different pathways to abnormal angiogenesis that differ depending on preeclampsia and bleeding outcomes.

This study is novel both in its use of the NuMoM2b public dataset which offers a large, well-characterized dataset with biomarker data and in its findings of strong relationships between placental angiogenic biomarkers and postpartum bleeding outcomes. Important limitations remain. There was no direct measure of blood loss or +/-PPH diagnosis in the dataset, just retained placenta and PPH requiring blood transfusion. While the latter makes it clear a hemorrhage took place, it means that hemorrhage not requiring a blood transfusion is not captured in this analysis. This could mean that our findings are more reflective of the physiology of severe hemorrhage. Transfusion for PPH is a relatively rare event, indicated by a range of outcomes include bleeding over 1500 mL and significant postpartum decreases in hemaglobin and hematocrit [46, 47]. This highlights the need for identifying and recording a broader range of bleeding outcomes to resolve underlying pathophysiology. Retained placenta is one of the most common proximate causes of postpartum hemorrhage, along with atony, and is more likely with prolonged third stage of labor, with manual removal being recommended within 10-30 minutes to prevent hemorrhage [48, 49]. Thus even if our findings are reflective directly of only the risk of retained placenta or severe hemorrhage, they are still relevant to understanding hemorrhage risk generally. The sample size of women who experienced either of these outcomes was only 67 total, or just 3.1%. A recent analysis of temporal trends in PPH rates in the National Inpatient Sample (n=76.7 million delivery hospitalizations) found that the rate rose from 2.7% in 2000 to 4.3% in 2019 [5]; the nuMoM2b study period spanned 2010-2015, suggesting that limiting to just cases of retained placenta and PPH requiring blood transfusion did effect the potential sample size for this study. Our sample over-represented adverse outcomes, including pre-eclampsia, small for gestational age, preterm birth, and stillbirth, which may mean our findings are more applicable to higher risk pregnancies. Another limitation is that our analyses focus only on the relationship between these biomarkers and blood loss, with additional nuance provided by preeclampsia status. For this important first step, we did not model the potential contributions of demographic or other pregnancy and labor variables to the relationship between angiogenic biomarkers and bleeding.

## Conclusion

Our findings present a novel approach to screening for PPH risk both separate from preeclampsia and as a potential complication of preeclampsia, by identifying target biomarkers and temporal windows to differentiate risk and thus course of treatment and management of labor. Many treatments are available for PPH once it starts, but there are limited approaches to identifying and mitigating the likelihood of excessive blood loss earlier in gestation, which could be invaluable in resource-limited settings and reduce the morbidity associated even with treated PPH [50]. The recent FDA-approved biomarker screening test for preeclampsia [51], which measures sFlt-1 and PlGF, is recommended for use between 23 and 35 weeks gestation. Our findings provide hope that someday we will be able to identify these and other biomarkers as reliable predictors of PPH risk, potentially even earlier in pregnancy, signifying a great change in our approach to the treatment and prevention of PPH.

## Data Availability

nuMoM2B data can be obtained through application to the National Institute of Child Health and Human Development Data and Specimen Hub.

https://dash.nichd.nih.gov/

## Ethics statement

Institutional Review Board (IRB) approval for the parent nuMoM2b study was obtained from Columbia University Human Subjects Institutional Review Board and the City University of New York (IRB-AAAR9413 and .HRPP/IRB 2019-0855 respectively). Each site reviewed and approved all study procedures. All participants (or their legal guardian(s)) provided written informed consent before participating in the study. All methods in the parent study and this secondary analysis were carried out in accordance with relevant guidelines and regulations. This secondary analysis of deidentified data is exempt from IRB approval beyond that granted to the parent study.

## Funding and acknowledgments

This study was funded by the University of Arizona College of Nursing (JNR, PI) and R00NR019596 (EE, PI; EG, Postdoctoral Research Fellow). The authors thank Elizabeth Abrams, Mary Dawn Koenig, Hillary Ruvalcaba, and Lily Woods. JNR thanks Adelaide Caledonia Goehl for her collaboration on a beautiful placenta.

## References

1. Shields, L., D. Goffman, and A. Caughey, Practice Bulletin No. 183: Postpartum Hemorrhage. Obstetrics & Gynecology, 2017. 130(4): p. e168–e186.

2. WHO, A roadmap to combat postpartum haemorrhage between 2023 and 2030. 2023, Geneva: World Health Organization. 68.

3. Tuncalp, O., et al., New WHO recommendations on prevention and treatment of postpartum hemorrhage. Int J Gynaecol Obstet, 2013. 123(3): p. 254–6.

4. Say, L., et al., Global causes of maternal death: a WHO systematic analysis. Lancet Global Health, 2014. 2(6): p. e323–333.

5. Corbetta-Rastelli, C., et al., Postpartum Hemorrhage Trends and Outcomes in the United States, 2000-2019. Obstetrics & Gynecology, 2023. 141(1): p. 152–161.

6. Ahmadzia, H., C. Grotegut, and A. James, A national update on rates of postpartum haemorrhage and related interventions. Blood Transfus, 2020. 18(4): p. 247–53.

7. Kramer, M., et al., Risk Factors for Postpartum Hemorrhage: Can We Explain the Recent Temporal Increase? J Obstet Gynaecol Can, 2011. 33(8): p. 810–819.

8. Ramanathan, G. and S. Arulkumaran, Postpartum hemorrhage. J Obstet Gynaecol Can, 2006. 28(11): p. 967–973.

9. Maughan, K.L., S.W. Heim, and S.S. Galazka, Preventing postpartum hemorrhage: managing the third stage of labor. Am Fam Physician, 2006. 73(6): p. 1025–8.

10. Prata, N., et al., Inability to predict postpartum hemorrhage: insights from Egyptian intervention data. BMC Pregnancy Childbirth, 2011. 11: p. 97.

11. ACOG, 10.1111/ajo.13599. Obstetrics & Gynecology, 2019. 134(6): p. e150–156.

12. Carr, B., et al., Predicting postpartum haemorrhage: a systematic review of prognostic models. Australia and New Zealand Journal of Obstetrics and Gynaecology, 2022. 62(6): p. 813–825.

13. Abrams, E.T. and J.N. Rutherford, Framing postpartum hemorrhage as a consequence of human placental biology: an evolutionary and comparative perspective. Am Anthropol, 2011. 113(3): p. 417–30.

14. Ruchob, R., J.N. Rutherford, and A.F. Bell, A Systematic Review of Placental Biomarkers Predicting Small-for-Gestational-Age Neonates. Biol Res Nurs, 2018. 20(3): p. 272–283.

15. Benschop, L., et al., Placental Growth Factor as an Indicator of Maternal Cardiovascular Risk After Pregnancy. Circulation, 2019. 139: p. 1698–1709.

16. Stangret, A., et al., Mild anemia during pregnancy upregulates placental vascularity development. Med Hypotheses, 2017. 102: p. 37–40.

17. Stangret, A., et al., Maternal hemoglobin concentration and hematocrit values may affect fetus development by influencing placental angiogenesis. J Matern Fetal Neonatal Med, 2017. 30(2): p. 199–204.

18. Cerdeira, A.S. and S.A. Karumanchi, Angiogenic factors in preeclampsia and related disorders. Cold Spring Harb Perspect Med, 2012. 2(11).

19. Zeisler, H., et al., Predictive Value of the sFlt-1:PlGF Ratio in Women with Suspected Preeclampsia. New England Journal of Medicine, 2016. 374: p. 13–22.

20. Buhimschi, C., et al., Urinary angiogenic factors cluster hypertensive disorders and identify women with severe preeclampsia. American Journal of Obstetrics and Gynecology, 2005. 192(3): p. 734–741.

21. Verlohren, S., et al., Clinical interpretation and implementation of the sFlt-1/PlGF ratio in the prediction, diagnosis and management of preeclampsia. Pregnancy Hypertension, 2022. 27: p. 42–50.

22. Maynard, S.E., et al., Excess placental soluble fms-like tyrosine kinase 1 (sFlt1) may contribute to endothelial dysfunction, hypertension, and proteinuria in preeclampsia. J Clin Invest, 2003. 111(5): p. 649–58.

23. Verlohren, S., et al., New Gestational Phase–Specific Cutoff Values for the Use of the Soluble fms-Like Tyrosine Kinase-1/Placental Growth Factor Ratio as a Diagnostic Test for Preeclampsia. Hypertension, 2013. 63(2): p. 346–352.

24. Levine, R., et al., Soluble Endoglin and Other Circulating Antiangiogenic Factors in Preeclampsia. New England Journal of Medicine, 2006. 355: p. 992–1005.

25. DeVivo, A., et al., Endoglin, PlGF and sFlt-1 as markers for predicting pre-eclampsia. Acta Obstetricia et Gynecologica Scandinavica, 2008. 87(8): p. 837–842.

26. Iannaccone, A., et al., Soluble endoglin versus sFlt-1/PlGF ratio: detection of preeclampsia, HELLP syndrome, and FGR in a high-risk cohort. Hypertension in Pregnancy, 2022. 41(3-4): p. 159–172.

27. Yan, X., R. Baxter, and S. Firth, Involvement of pregnancy-associated plasma protein-A2 in insulin-like growth factor (IGF) binding protein-5 proteolysis during pregnancy: a potential mechanism for increasing IGF bioavailability. Journal of Clinical Endocrinology & Metabolism, 2010. 95: p. 1412–1420.

28. Luewan, S., et al., Low maternal serum pregnancy-associated plasma protein-A as a risk factor of preeclampsia. Singapore Medical Journal, 2018. 59(1): p. 55–59.

29. Neuman, R., et al., PAPPLA2 and Inhibin A as Novel Predictors for Pregnancy Complications in Women With Suspected or Confirmed Preeclampsia. Journal of the American Heart Association, 2020. 29(9): p. e018219.

30. Ree, P., et al., Early detection of preeclampsia using inhibin a and other second-trimester serum markers. Fetal Diagnosis and Therapy, 2011. 29(4): p. 280–286.

31. Herr, F., et al., HCG in the regulation of placental angiogenesis. Results of an in vitro study. Placenta, 2007. 28 p. 85–93.

32. Sharony, R., et al., The association between maternal serum first trimester free βhCG, second trimester intact hCG levels and foetal growth restriction and preeclampsia. Journal of Obstetrics and Gynaecology, 2018. 38(3): p. 363–366.

33. Zhang, X., et al., Predictive Performance of Serum β-hCG MoM Levels for Preeclampsia Screening: A Meta-Analysis. Frontiers in Endocrinology, 2021. 12: p. 619530.

34. Skogler, J., et al., Association between human chorionic gonadotropin (hCG) levels and adverse pregnancy outcomes: A systematic review and meta-analysis. Pregnancy Hypertension, 2023. 34: p. 124–137.

35. Abrams, E. and J. Rutherford, Is postpartum hemorrhage a legacy of our evolutionary past?, in A Textbook of Postpartum Hemorrhage, L. Keith and C. Jones, Editors. 2012, Sapiens: London. p. 55–63.

36. Farina, A., et al., Elevated maternal placental protein 13 serum levels at term of pregnancy in postpartum major hemorrhage (>1000 mLs). A prospective cohort study. American Journal of Reproductive Immunology, 2017. 78: p. e12702.

37. Haas, D., et al., A description of the methods of the Nulliparous Pregnancy Outcomes Study: monitoring mothers-to-be (nuMoM2b). American Journal of Obstetrics and Gynecology, 2015. 212(4): p. 539.E1-539.E24.

38. Silver, R., Nulliparous Pregnancy Outcomes Study: Monitoring Mothers-to-be [dataset], N.D.a.S. Hub, Editor. 2019.

39. Eskild, A., et al., Levels of angiogenic factors in pregnancy and post-partum bleeding. Acta Obstetrica et Gynecologica Scandinavica, 2010. 87(10): p. 1081–1083.

40. Ghosh, S.K., et al., Association between placental growth factor levels in early onset preeclampsia with the occurrence of postpartum hemorrhage: A prospective cohort study. Pregnancy Hypertens, 2012. 2(2): p. 115–22.

41. Cai, G. and S. Zhao, Correlation analysis of AFP and β-hCG on obstetric complications and pregnancy outcomes. International Journal of Clinical and Experimental Medicins, 2020. 13(8): p. 6026–6031.

42. Chen, Y., et al., Relationship between increased maternal serum free human chorionic gonadotropin levels in the second trimester and adverse pregnancy outcomes: a retrospective cohort study. BMC Womens Health, 2024. 24: p. 323.

43. Aquilina, J., et al., Second-trimester maternal serum inhibin A concentration as an early marker for preeclampsia. American Journal of Obstetrics and Gynecology, 1999. 181(1): p. 131–136.

44. Singnoi, W., et al., A cohort study of the association between maternal serum Inhibin-A and adverse pregnancy outcomes: a population-based study. BMC Pregnancy Childbirth, 2019. 19: p. 124.

45. Balkas, G. and T. Caglar, Elevated first-trimester PAPP-A is a marker in high-risk pregnancies with an increased risk of placenta accreta in predicting adverse outcomes. European Review for Medical and Pharmacological Sciences, 2023. 27(20): p. 9955–9961.

46. Ruiz-Labarta, F., et al., Red Blood Cell Transfusion after Postpartum Hemorrhage: Clinical Variables Associated with Lack of Postpartum Hemorrhage Etiology Identification. Journal of Clinical Medicine, 2023. 12(19): p. 6175.

47. WHO, WHO recommendations for the prevention and treatment of postpartum haemorrhage. 2012.

48. Chikkamath, S., et al., Duration of third stage labour and postpartum blood loss: a secondary analysis of the WHO CHAMPION trial data. Reproductive Health, 2021. 2021: p. 230.

49. Frolova, A., et al., Duration of the Third Stage of Labor and Risk of Postpartum Hemorrhage. Obstetrics & Gynecology, 2016. 127(951-956).

50. Doumouchtsis, S., A. Papageorghiou, and S. Arulkumaran, Systematic Review of Conservative Management of Postpartum Hemorrhage: What to Do When Medical Treatment Fails. Obstetrical & Gynecological Survey, 2007. 62(8): p. 540–547.

51. Hopkins Tanne, J., Pre-eclampsia: FDA approves blood test to identify pregnant women at risk. British Journal of Medicine, 2023. 382: p. 1594.

